# Machine learning models aimed at identifying risk factors for reducing morbidity and mortality still need to consider confounding related to calendar time variations

**DOI:** 10.1101/2022.05.24.22275482

**Authors:** Andreas Rieckmann, Tri-Long Nguyen, Piotr Dworzynski, Ane Bærent Fisker, Naja Hulvej Rod, Claus Thorn Ekstrøm

**Affiliations:** Section of Epidemiology, Department of Public Health, University of Copenhagen, Denmark; Novo Nordisk Foundation Center for Basic Metabolic Research, University of Copenhagen, Denmark; OPEN, University of Southern Denmark, Odense, Denmark; Indepth Network, Bandim Health Project, Bissau, Guinea-Bissau; Section of Biostatistics, Department of Public Health, University of Copenhagen, Denmark

**Keywords:** Machine learning, Causal inference, Confounding, Bias, Health Data

## Abstract

Machine learning models applied to health data may help health professionals to prioritize resources by identifying risk factors that may reduce morbidity and mortality. However, many novel machine learning papers on this topic neither account for nor discuss biases due to calendar time variations. Often, efforts to account for calendar time (among other confounders) are necessary since patterns in health data – especially in low- and middle-income countries – may be influenced by calendar time variations such as temporal changes in risk factors and changes in the disease and mortality distributions over time (epidemiological transitions), seasonal changes in risk factors and disease and mortality distributions, as well as co-occurring artefacts in data due to changes in surveillance and diagnostics. Based on simulations, real-life data from Guinea-Bissau, and examples drawn from recent studies, we discuss how including calendar time variations in machine learning models is beneficial for generating more relevant and actionable results. In this brief report, we stress that explicitly handling temporal structures in machine learning models still remains to be considered (like in general epidemiological studies) to prevent resources from being misdirected to ineffective interventions.

## Introduction

The introduction of machine learning (ML) models into public health analyses brought a promise of aiding the reduction in mortality and morbidity by making predictions (e.g. identifying individuals at high risk) or increasing our understanding of disease patterns by uncovering risk factors (e.g. why an individual is at risk).^1^ Though ML models may also be used to reduce high-dimensional datasets, causal estimation, and to identify unfair biases hidden in data, studies on prediction and the risk factors important for the prediction are by far the majority.^2^ Although the term “*risk factors”* avoids making claims about causality, researchers need to consider causes and effects to develop interventions for improving health.^3–5^ This concerns both: risk factors to be intervened upon and risk factors used to identify high-risk groups.

Many novel ML papers identifying risk factors do not account for or discuss calendar time variations.^6–13^ This is not necessarily problematic, but it can result in misleading conclusions. This is relevant to all national public health strategies, but especially to low- and middle-income countries (LMIC), since some may struggle with a double burden of disease. The changes in the disease distributions over time from communicable diseases to non-communicable diseases occur at such a fast pace that the health system has to accommodate a more complex patient case-mix than in countries where non-communicable diseases are dominant (accelerated epidemiological transition).^14^ In only 25 years (between 1990 and 2015) child mortality declined by 54% in Sub-Saharan Africa^15^, and the burden of communicable diseases continued to decrease.^16^ In this sense, long-term health data from LMIC countries in accelerated epidemiological transitions may be confounded by temporal structures if some of the risk factors of interest are also affected by temporal structures (e.g. due to economic development). Thus, the identification of risk factors may be distorted by calendar time, and to adjust for this type of confounding, calendar time must be included in the ML model. Also, seasonal changes may affect a variety of risk factors as well as diseases and mortality in short-term studies, and thus an operationalization of seasonality must be included in the ML model. Finally, spurious correlations can also occur simply due to co-occurring changes in surveillance and diagnostic criteria. Simple robustness checks of whether the inclusion of calendar time variations affects the results can help direct the researchers to a better understanding of the studied phenomenon.

## Materials and methods

This paper briefly summarizes the phenomenon confounding from the causal inference literature to give two examples of how calendar time variations may misdirect us in a simulated data example and an example of real data from Guinea-Bissau. In the Discussion, we relate the issue to published scientific literature.

### A summary of confounding

The causal inference literature formalizes questions that deal with quantifying the effect of a well-defined intervention on a health outcome.^17^ A key approach to understanding causal relationships is the study of causal structures. Directed acyclic graphs (DAG) depict causal structures and help researchers to decide which variables are necessary to adjust for. In a DAG, causal structures between features are shown as single-headed arrows. In Figure 1A, the arrow X→ Y denotes a causal relation: if X changes, Y also changes; but if Y changes, X does not change. In Figure 1B, the structure X ←C→Y means that if C changes then both X and Y change. In data where C is not observed, X and Y are correlated since they share a common (unobserved) cause C. However, X has no causal effect on Y nor does Y have a causal effect on X – their relationship is *confounded* by C, the common, underlying cause. To prevent X and Y from being spuriously associated, feature C needs to be adjusted in the model.

**Figure 1.**
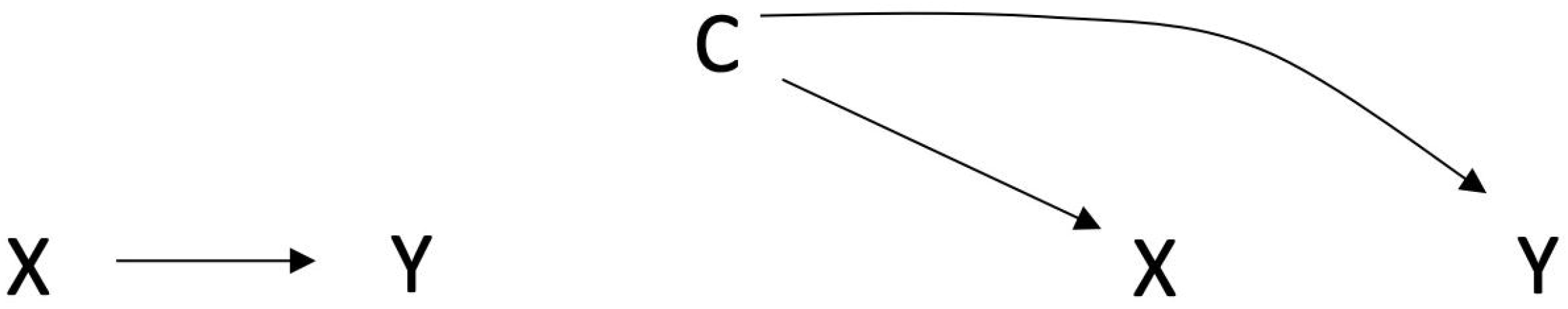
Directed Acyclic Graphs. A) A direct effect of X on Y B) A spurious association between X and Y due to confounding

### Examples

To illustrate the importance of including calendar time variations as a potential confounder in machine learning algorithms, we considered two situations: simulated data where we knew the exact data-generating structure and a real-life application on data from rural Guinea-Bissau. As one example of an ML model, we employed a single hidden layer neural network with 5 hidden neurons, which took all inputs as one-hot-encoded features. The activation functions were identical for each neuron and output layer but were chosen specifically for each case as explained below.

We implemented the analyses in R^18^ using Keras^19^ and Tensorflow^20^ frameworks. R script and performance curves for the simulation can be found in Supplementary code 1 and Supplementary figure 1.

### Simulation

Figure 2 shows the causal DAG for a simple data structure of binary features, where calendar time influences two features (X_2_ and X_12_) along with the outcome, Y. X_10_ has an effect on the outcome itself. We used this structure to simulate data sets of 10,000 individuals for training, 5,000 individuals for validation, and 10,000 individuals for out-of-sample evaluation. We fit two neural networks – one with and one without calendar time - with logistic sigmoid activation functions. Using stochastic gradient descent with batches of 10, training was stopped at the first epoch with an increase in loss with a patience of 10 epochs on the test data set. We estimated the variable importance by permuting (/randomizing the order of) one feature at a time of the out-of-sample evaluation data and compared the difference in the area under the receiver operating characteristic curve (ROC AUC) from the true ROC AUC using out-of-sample validation data. For each simulation, the mean of the decrease in ROC AUC from 10 permutations of each variable was saved. We reported estimates from 100 of the above simulations in box plots for the models with and without calendar time.

**Figure 2.**
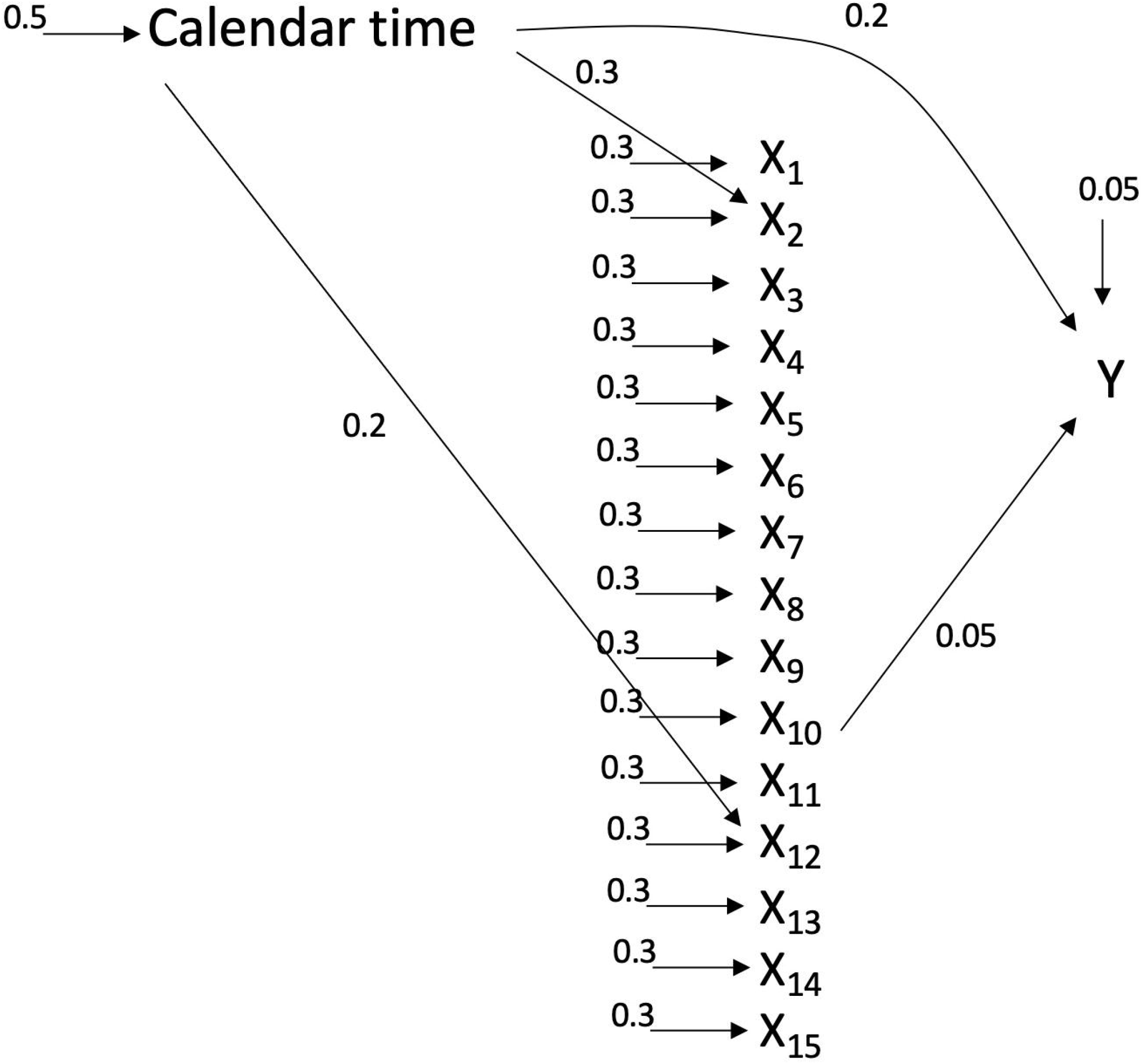
Data generation set up for simulation. Calendar time acts as a confounder for X2 and X12. Numerical values indicate the additive probability of each binary variable. X_1-15_ are all inputs. Y is the outcome. Calendar time influences X_2_ and X_12_. X_10_ influences Y.

### Real-life data from Guinea-Bissau

To demonstrate our point in real data, we used data from the rural areas of Guinea-Bissau, West Africa, gathered as part of the Bandim Health Project (BHP), which runs a longitudinal health and demographic surveillance system (HDSS).^21^ We included 29,609 children from 182 village clusters in 10 regions under surveillance between the 1^st^ of June 2008 and the 31^st^ of May 2011. Villages are visited biannually by mobile teams and vital information is recorded for children, preferably from pregnancy, to their fifth birthday. We analyzed this cohort of children in monthly intervals for whether a child had died or not. Children emigrating would be censored from the forthcoming month. We included information about sex, education of the primary caretaker [none, 1-4 years, 5+ years, missing], and age of the primary caretaker [10-19 years, 20-29 years, 30-39 years, 40-49 years, 50+ years, missing]. In this analysis, missing values were given its own category, but researchers could argue for restricting the population to individuals with full information (if a missing completely-at-random situation can be assumed) or imputing the missing values if the reason for having missing values depends on underlying structures related with the exposures and the outcome (and the missing data are missing-at-random).^22^ Furthermore, we created a variable representing a hypothetical intervention, say a nationwide distribution of insecticide-treated nets for malaria prevention, on the 1^st^ of December 2009, to which all children were “exposed” thereafter. In the adjusted model, we also included a seasonality variable coded as rainy season from the 1^st^ of June to the 30^th^ of November, and as dry season from the 1^st^ of December to the 30^th^ of May.

Due to computational challenges as a result of the expansion of the dataset from 29,609 children to 591,020 child observation months, we analysed a subset of data consisting of all observation months ending with a death (1384) and a random sample of 20,000 observation months ending in no death. By outcome value, this data was split into 33% for an out-of-sample evaluation data set. The remaining data for training the models were further split into 66% for training data and 33% for test data for early stopping stratified by the outcome value.

We fit two neural networks – one without calendar time and one with calendar time - with rectifier linear units (ReLU) activation functions. We initiated all weights with positive values close to 0, however, the bias to the output layer was initiated with the proportion of child observation months with a death in the analyzed data. Using stochastic gradient descent with batches of 1, training was stopped at the first epoch with an increase in loss with a patience of 5 epochs on the test data set. When training, observation periods were weighted with the inverse of the prevalence of the observation’s outcome value in the training dataset. We estimated the variable importance by permuting (/randomizing the order of) one feature at a time of the out-of-sample evaluation data and compared the difference in the ROC AUC from the ROC AUC using the non-permuted out-of-sample validation data. For each simulation, the mean of the decrease in ROC AUC from 50 permutations of each variable was saved. We report estimates from 100 of the above simulations in box plots for each model with and without seasonality.

## Results

### Simulation

Figure 3 shows that X_2_, X_10_, and X_12_ all were highly important for the model when calendar time was not included. We observed that the spurious associations with X_2_ and X_12_ disappeared when we took calendar time into account by including the variable as an input feature in the neural network. We also found that among the X-features, X_10_ was the only important factor influencing the outcome Y after including the calendar time variable in the model (Figure 3). Thus, including calendar time helped us to avoid falsely identifying risk factors for potential interventions, which did not causally affect the outcome.

**Figure 3.**
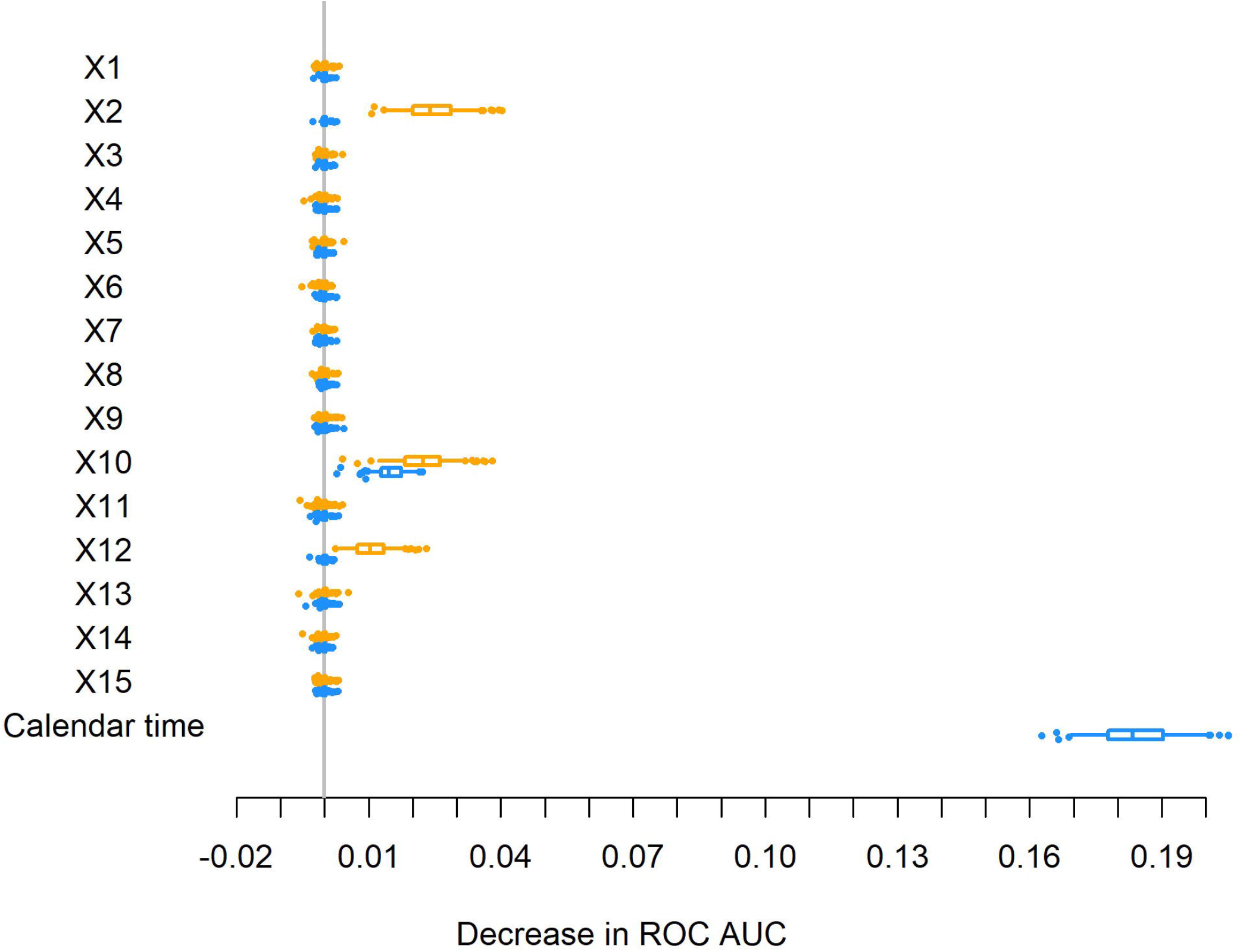
Variable importance in a neural network with and without calendar time in the simulation example. 100 estimations of variable importance measures from two one-hidden layer neural networks, of which one did not include information about calendar time (Orange), and another which included information about calendar time (Blue). Only variable X_10_ has a real causal effect on the outcome. If calendar time is not included then features sharing a common cause with the outcome, X_2_ and X_12_, are spuriously presented as important variables.

### Real-life data from Guinea-Bissau

Our results from both models indicate that the education of the primary caretaker is an important feature for predicting child mortality among the included risk factors in this time period (Figure 4). The variable importance measures for sex, education and age of the primary caretaker are very similar in the model with and without seasonality. The hypothetical intervention had an importance measure similar to the size of education of the primary caretaker in the model without seasonality, however, the importance of the hypothetical intervention dropped when seasonality was introduced in the model. This suggests that the result related to the hypothetical intervention is sensitive to time changes such as seasonality, and may require further investigation by the researchers.

**Figure 4.**
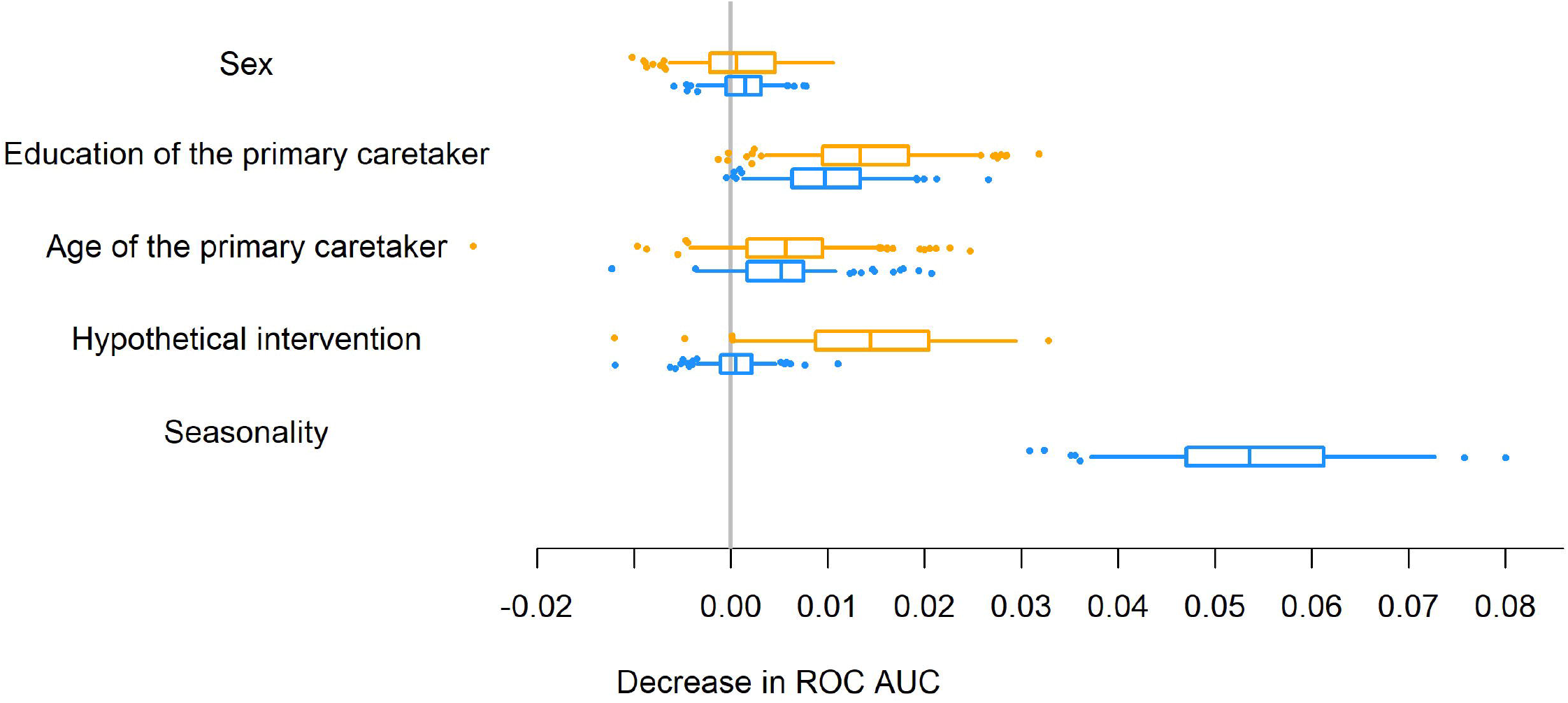
Variable importance in a neural network with and without seasonality in the real-life data example. 100 estimations of variable importance measures from two one-hidden layer neural networks, of which one did not include information about seasonality (Orange), and another which included information about seasonality (Blue). The variable importance of the hypothetical interventions drops when the season is included in the model.

The monthly mortality rate follows sine curve fluctuations throughout the years (Supplementary Figure 2), with almost twice the mortality in the rainy season compared with the mortality in the dry season. It becomes evident that the time period before the hypothetical intervention has two rainy seasons and one dry season, while the time period after the hypothetical intervention has two dry seasons and one rainy season, which naturally should be accounted for before interpreting the importance of the hypothetical intervention.

## Discussion

We have demonstrated the reasoning behind and use of including operationalization of calendar time variations in machine learning models. We highlight that this understanding can also be used as a robustness check on whether the variable importance measures are sensitive to the inclusion of confounding variables such as calendar time variations.

We give two examples of how this concern applies to current research using ML models for public health goals. In an ML-aided study published in 2017, Tuti *et al*. studied more than 10 000 children, aged 2-59 months, admitted with clinical non-severe pneumonia at 14 hospitals in Kenya between February 2014 and February 2016.^6^ The authors identified risk factors for inpatient mortality using five models and found that comorbidity (measured as a diagnosis of malaria, diarrhoea, dehydration or anaemia) was in average the fifth most important of the included risk factors. We do not attempt to challenge that children with comorbidities are more susceptible to causes of pneumonia or have a higher case-fatality rate. However, given that malaria incidence and pneumonia mortality follow seasonal patterns in Kenya,^23,24^ adjusting for calendar time and seasonality (by including them as input features) would have supported the authors’ conclusion on the importance of the comorbidities. In another ML-aided study published in 2018, Sauer *et al*. investigated risk factors for treatment failure among tuberculosis patients in LMIC.^7^ They used data from 587 tuberculosis patients from 2000 to 2016 in Azerbaijan, Georgia, Republic of Moldova, Romania and Belarus using seven models. They found that the type of drug-resistant tuberculosis and education were some of the most important risk factors. However, during the 16 years, the incidence of drug-resistant tuberculosis has changed world wide^25^ (as described by the authors^7^); tertiary education increased before 2012 in Belarus^26^; and the risk of tuberculosis treatment failure may have varied. Again, including calendar time in the model would have strengthened the conclusion about the relative importance of risk factors, for the aim of monitoring patient sub-groups at increased risk of treatment failure.

Thus, the inclusion of calendar time in ML models should still be carefully considered and constitutes a crucial step towards more relevant and actionable results, when risk factors are identified as potential areas for intervention. This is natural in epidemiological studies, but the literature suggests that it should be reminded in ML-based studies on health data. Issues stemming from the lack of inclusion of calendar time in ML models are not restricted to analyses of LMIC health data, but the problem may be exacerbated in data from countries with fast changes in risk factors and disease distributions over time. Furthermore, calendar time is often not the only confounder which misdirects emphasis on irrelevant risk factors, and other underlying common causes should be identified and controlled for before drawing causal conclusions. Additionally, spurious correlations may also be introduced by including common effects or mediators in the model. Relevant literature about the identification of risk factors under a causal framework might be of great interest to researchers engaged in improving their ML-based healthcare models for actionable results.^3–5^

## Conclusions

In conclusion, our paper is a reminder for researchers conducting ML-based studies on individual-level health data, that it remains essential to explicitly handle confounder structures such as calendar time variations when identifying risk factors and disease patterns. Including calendar time and seasonality may help researchers distinguish risk factors from general trends, and thus suggest more relevant health interventions to improve public health.

## Data Availability

The simulated dataset can be generated using the R script in the supplementary material. Request for data access is referred to Bandim Health Project.

## Acknowledgement

The analysis of real-life data, would not have been possible without the dedicated work of the many data collectors and supervisors, as well as mothers of children in the villages under surveillance who were willing to provide an answer to the questions.

## Ethics statement

The BHP HDSS surveillance was initiated in 1990 at the request of the Ministry of Health. Surveyed women provided oral consent at the time of registration. Protocols for concurrent trials nested in the HDSS and describing the data collection have been approved by the Ministry of Health (Núcleo de Coordencão das Pesquisas do Ministério da Saúde: NPC no. 12/2007, NPC no. 02/2008), National Ethics Committee in Guinea-Bissau (Comite Nacional de Etica na Saude: no 34/CNES/2010, 08/CNES/2011) and received consultative approval from the Central Ethical Committee in Denmark (2006-7041-99; 1103988)

## Author contributions

All authors made substantial contributions to the conception of the work. AR conducted the literature search and carried out the simulation and real-life data analyses. ABF guided the conceptualization of the real-life data example with an understanding of the context and data gathering process. AR and TLN drafted the manuscript, and PD, ABF, NHR, and CTE critically revised it for important intellectual content.

## Competing interests

None.

## Funding

AR was supported by an international postdoc grant by the Independent Research Fund Denmark (9034-00006B). PD was supported by a research grant from the Danish Diabetes Academy funded by the Novo Nordisk Foundation.

## Notes

### Competing Interest Statement

The authors have declared no competing interest.

### Author Declarations

The BHP HDSS surveillance was initiated in 1990 at the request of the Ministry of Health. Surveyed women provided oral consent at the time of registration. Protocols for concurrent trials nested in the HDSS and describing the data collection have been approved by the Ministry of Health (Nucleo de Coordencao das Pesquisas do Ministerio da Saude: NPC no. 12/2007, NPC no. 02/2008), National Ethics Committee in Guinea-Bissau (Comite Nacional de Etica na Saude: no 34/CNES/2010, 08/CNES/2011) and received consultative approval from the Central Ethical Committee in Denmark (2006-7041-99; 1103988)

