## supplementary material for "Machine learning models aimed at identifying risk factors for reducing morbidity and mortality still need to consider confounding related to calendar time variations"

### Supplementary code 1. R script for simulations

```
################################## Defining functions ##################################

# Simulated example
library(pROC)
library(keras)

gen_data <- function(n) {
data <- data.frame(V1 = sample(0:1,n,replace = T))
for (i in 1:15) {
  data[,i] <- sample(0:1,n,replace = T, prob = c(0.7,0.3))
}
summary(data)

C = as.numeric(sample(0:1,n,replace=T,prob = c(0.5,0.5)))

for (i in 1:nrow(data)) {
  if (C[i]==1 & sample(0:1,1,prob = c(0.7,0.3))==1) {
    data[i,2] <- 1
  } }

for (i in 1:nrow(data)) {
  if (C[i]==1 & sample(0:1,1,prob = c(0.8,0.2))==1) {
    data[i,12] <- 1
  } }

data$Y <-  sample(0:1,n,replace = T, prob = c(0.95,0.05))

for (i in 1:nrow(data)) {
  if (C[i]==1 & sample(0:1,1,prob = c(0.8,0.2))==1) {
    data$Y[i] <- 1
  } }
for (i in 1:nrow(data)) {
  if (data[i,10]==1 & sample(0:1,1,prob = c(0.95,0.05))==1) {
    data$Y[i] <- 1
  } }

for(i in 1:ncol(data)) {
  data[,i] <- as.numeric(data[,i])
}

data$C <- C
data <- data[,c(16,1:15,17)]

return(data)
}

importance<- function(y,data, model,nsim=10) {
  ref <- auc(y,as.vector(predict(model, as.matrix(data))))
  aucs <- matrix(0,nrow=nsim,ncol=ncol(data))
  for (i in 1:ncol(data)) {
    print(i)
    xs <- data
    for (g in 1:nsim) {
      xs[,i] <- xs[sample(1:nrow(xs)),i] # permute
    aucs[g,i] <- ref-auc(y,as.vector(predict(model, as.matrix(xs))))
    }
  }
  aucs <- data.frame(aucs)
  colnames(aucs) <- names(data)
  return(aucs)
  }

plot_importance <- function(results){
  #c(min(results)-0.2*diff(range(results)),max(results))
  plot(0,ylim=c(0,16),xlim=c(-0.07,0.05),xlab="Decrease in AUC",ylab="",main="Variable importance",
```

```
      axes=F)
  axis(1,seq(-0.02,0.1,0.01))
  abline(v=0,lty=1,col="grey",lwd=2)
  for(i in ncol(results):1) {
    text(-0.05,16-i, bquote(X[.(i)])) #names(results)[i])
    arrows(x0=mean(results[,i]),x1=quantile(results[,i],0.975),y0=16-i,angle=90,length=0.05)
    arrows(x0=mean(results[,i]),x1=quantile(results[,i],0.025),y0=16-i,angle=90,length=0.05)
    points(mean(results[,i]),16-i,pch=16,col=adjustcolor("black",1))
  }
}

results_1_1 <- data.frame(matrix(0,nrow=0,ncol=15))
names_1 <- names(results_1_1)
results_1_2 <- data.frame(matrix(0,nrow=0,ncol=16))
names_2 <- names(results_1_2)

for (run in 1:9) {
print(paste0("Simulation number: ",run))

#### Simulated data #####
n_sim = 10
set_batch_size = 10
set_epochs = 2000
set_nodes = 5

data <- gen_data(15000)
y <- data[,1]
data <- data[,-1]
data_no_c <- data[,-ncol(data)]
c <- data[,ncol(data)]

test_data <- gen_data(10000)
test_y <- test_data[,1]
test_data <- test_data[,-1]
test_data_no_c <- test_data[,-ncol(test_data)]
test_c <- test_data[,ncol(test_data)]


################ A) Variable importance in a neural network without calendar time #################
inputs <- layer_input(shape = ncol(data_no_c))
predictions <- inputs %>%
  layer_dense(units = set_nodes, activation = 'sigmoid') %>%
  layer_dense(units = 1, activation = 'sigmoid')
model <- keras_model(inputs = inputs, outputs = predictions)
summary(model)
model %>% compile(optimizer = 'sgd', loss = 'mean_squared_error', metrics = c('accuracy'))
early_stop <- callback_early_stopping(monitor = "val_loss", patience = 10, restore_best_weights = TRUE)

history <- fit(
  object           = model,
  x                = as.matrix(data_no_c),
  y                = y,
  batch_size       = set_batch_size,
  epochs           = set_epochs,
  validation_split = 0.33333,
  callbacks        =early_stop
)
permutations_1 <- importance(test_y,test_data_no_c,model, nsim = n_sim)
results_1_1 <- rbind(results_1_1,colMeans(permutations_1))
performance_1 <- history$metrics$loss #Saved for the last simulation
performance_val_1 <- history$metrics$val_loss #Saved for the last simulation

################ B) Variable importance in a neural network with calendar time #################
inputs <- layer_input(shape = ncol(data))
predictions <- inputs %>%
  layer_dense(units = set_nodes, activation = 'sigmoid') %>%
  layer_dense(units = 1, activation = 'sigmoid')
model <- keras_model(inputs = inputs, outputs = predictions)
summary(model)
model %>% compile(optimizer = 'sgd', loss = 'mean_squared_error', metrics = c('accuracy'))
early_stop <- callback_early_stopping(monitor = "val_loss", patience = 10, restore_best_weights = TRUE)
```

```
history <- fit(
  object           = model,
  x                = as.matrix(data),
  y                = y,
  batch_size       = set_batch_size,
  epochs           = set_epochs,
  validation_split = 0.33333,
  callbacks        =early_stop
)
permutations_2 <- importance(test_y,test_data,model, nsim = n_sim)
results_1_2 <- rbind(results_1_2,colMeans(permutations_2))
performance_2 <- history$metrics$loss #Saved for the last simulation
performance_val_2 <- history$metrics$val_loss #Saved for the last simulation

save.image("data_sim.RData")

}

######################################################################
#Figure 3. A) Variable importance in a neural network without calendar time B) Variable importance in a  neural
network with calendar time
par(mfrow=c(1,2))
colnames(results_1_1) <- names_1
colnames(results_1_2) <- names_2
plot_importance(results_1_1)
plot_importance(results_1_2)
######################################################################
# Supplementary figure 1. Performance curve for a model without and with calendar time for one simulation
par(mfrow=c(1,2))
plot(performance_1,type='l',xlab="Epochs",ylab="Validation mean squared error",main="Performance curve for a
model\nwithout calendar time for one simulation",
     ylim=range(c(performance_1,performance_val_1)))
points(performance_val_1,type="l",lty=3,lwd=4)
plot(performance_2,type='l',lty=1,xlab="Epochs",ylab="Validation mean squared error",main="Performance curve
for a model\nwith calendar time for one simulation",
     ylim=range(c(performance_2,performance_val_2)))
points(performance_val_2,type="l",lty=3,lwd=4)


################################# End #####################################
```

**Supplementary figure 1. Performance curve for a model without and with calendar time for one simulation**

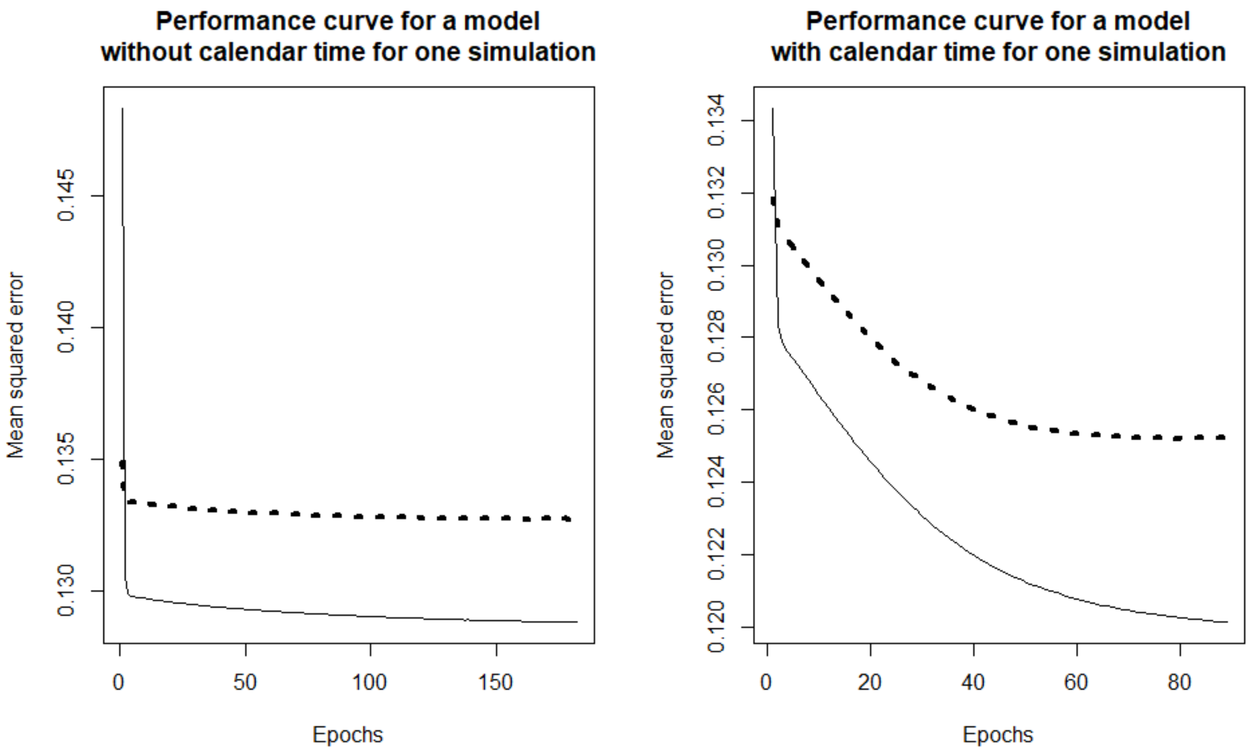


*The full line represents the training performance, the dotted line represents the test performance.*

**Supplementary figure 2. Seasonal differences in under 5 mortality in rural Guinea-Bissau during follow-up**

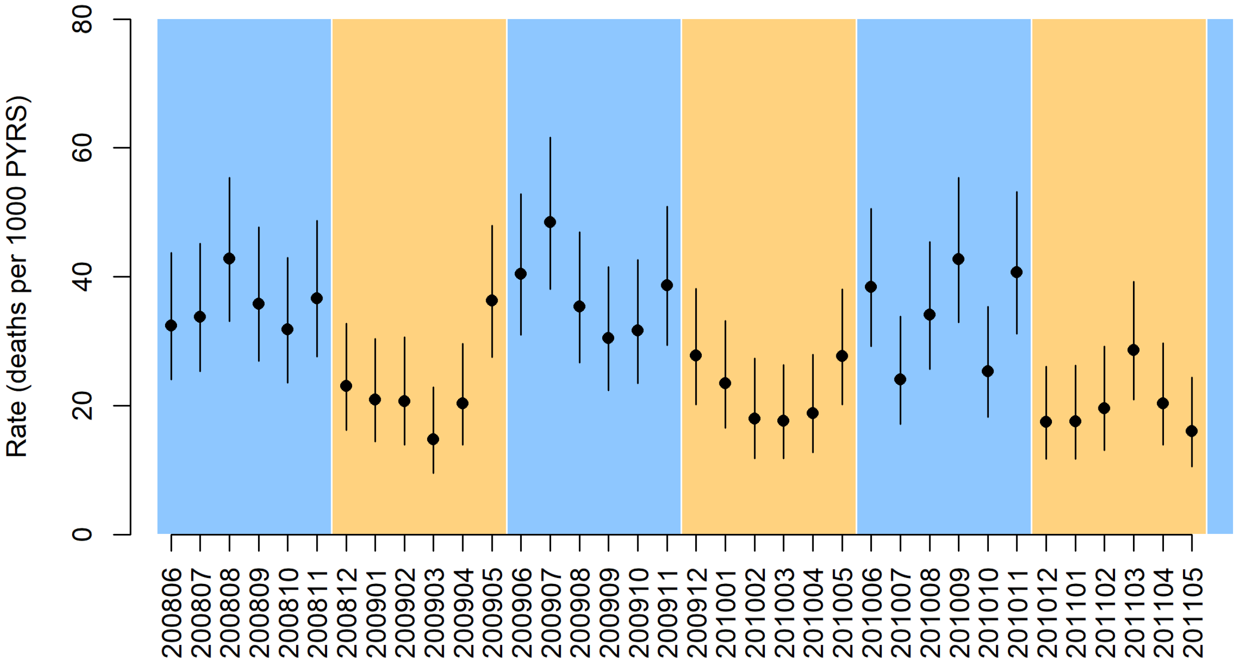


*The under 5 mortality rate by month and year. Blue time periods indicate the rainy season, orange time periods indicate the dry season.*
